# A Patient-Reported Measure of Locomotor Function Derived from the Functional Assessment Questionnaire

**DOI:** 10.1101/2021.06.12.21258826

**Authors:** Michael H. Schwartz, Nanette Aldahondo, Bruce A. MacWilliams

## Abstract

**Background:** Locomotor function is often impaired in children diagnosed with cerebral palsy (CP). Improving locomotor function is a common goal of treatment. The current gold standard for assessing locomotor function in CP is the gross motor function measure (GMFM-66). The GMFM-66 requires an in-person assessment by a trained clinician. It would be useful to have a measure of function that is like the GMFM-66 but can be assessed through patient report.

**Methods:** We queried the clinical databases of two motion analysis centers (Gillette Children’s Specialty Healthcare and Shriners Hospital – Salt Lake City) for individuals with a diagnosis of cerebral palsy (CP) who were 18 years old or younger and had undergone instrumented clinical gait analysis that included the functional assessment questionnaire (FAQ). We computed the transformed FAQ (FAQt) as the weighted sum of the skills an individual was able to perform, where the weighting was the difficulty of the skills. We assessed concurrent and external validity of the FAQt by comparing it to the GMFM-66.

**Results:** The FAQt exhibited strong concurrent and external validity. Linear regression showed that the GMFM-66 explained 54% of the variance in FAQt, and the linear fit was independent of center. The FAQt evolved with age in a manner similar to the GMFM-66, with higher functioning individuals, as measured by gross motor function classification system level, achieving higher levels of function at a higher rate and an earlier age compared to their lower functioning peers. The findings with respect to GMFM-66 did not depend on the center at which the data was acquired.

**Conclusions:** The FAQt demonstrates strong concurrent and external validity, making it a useful measure of locomotor function.

## 1 Background

The primary goal of this study was to develop a simple, relatively fine-grained measure of locomotor function that is based on patient reported skill ability. Specifically, we defined the functional assessment questionnaire transform (FAQt) as a weighted sum of overall walking ability and ability to perform 22 complex locomotor skills, as reported on the FAQ questionnaire. We showed that this measure exhibited strong concurrent validity compared to the gross motor function measure (GMFM-66) and external validity across two centers.

### 1.1 The Importance of Locomotor Function

Locomotor function is important for independence, recreation, participation in the community, health, and overall quality of life (1,2). For children diagnosed with cerebral palsy (CP) locomotor function is often impaired by primary neurological deficits (e.g., spasticity, weakness, poor motor control) and their orthopedic sequalae (e.g., excessive long bone torsion, muscle contracture). Improving locomotor function is one of goals of treatment for children with CP. By measuring locomotor function we can enhance our ability to assess impairments at the activity and participation level and better quantify the impact of treatments. There are several excellent, widely used measures of function already in existence, including the current gold-standard GMFM-66 (3).

### 1.2 The GMFM-66

The GMFM-66 is a task-based standardized measure administered by a physical therapist trained in its application. The test consists of five domains. The final two domains consist of the most difficult tests, evaluating standing (GMFM-D) and walking, running and jumping (GMFM-E). For ambulatory children it is typical that only these sections are administered since these individuals are expected to be able to complete all tasks in the other domains (A: Lying and Rolling, B: Sitting, and C: Crawling and Kneeling). Raw scores from sections D and E can be entered into a scoring database (Gross Motor Ability Estimator or GMAE) to compute a score normed to the full 66 items (GMFM-66) (4).

Despite its clear value, a significant limitation of the GMFM-66 is that it requires an in-person assessment by a trained clinician, which takes minutes approximately 20 minutes to administer (sections D and E only) and another few minutes to score using a digital database tool. This makes the GMFM-66 less practical for large studies, frequently repeated measurements, and patients with limited access to specialty healthcare services.

### 1.3 The FAQ

Our goal was to develop a measure of locomotor function that was granular, patient-reported, simple to assess, and easy to understand. We chose to start with an existing tool, then design modifications to make it suitable for our needs. The Functional Assessment Questionnaire (FAQ) was introduced over 20 years ago as a patient reported evaluation of mobility in children (5). We chose to start with the FAQ since it has been extensively validated, is well-established, free, and relatively widely used in practice. In addition, the walking and complex locomotor skills reported in the FAQ have been ranked for difficulty, and shown reflect a unimodal construct labeled “locomotor function” (6,7).

The FAQ consists of a single overall walking ability and 22 skill performance ratings, yielding 23 separate numbers. While these give a detailed picture of a patient’s locomotor function, they are difficult to distill and interpret globally. We have previously shown that, using the random forest algorithm, these 23 scores can be used to predict GMFCS level with around 90% accuracy, suggesting that there is strong concurrence between the locomotor function assessments underlying the FAQ and those reflected in the GMFCS (8). In the present study, we will derive a simple summary score from the FAQ and demonstrate compelling evidence of validity compared to the GMFM-66.

## 2 Methods

Approval for this study was granted by the University of Minnesota (STUDY00012420) and Western (STUDY1249365) institutional review boards.

### 2.1 Participants and Data

This study included data from the gait analysis services of two centers: Gillette Children’s Specialty Healthcare (GIL) and Shriners Hospital – Salt Lake City (SLC). We searched the databases of the centers for patients meeting the following criteria:

- Diagnosis of CP
- Up to 18 years old
- Gait assessment that included responses to the FAQ questionnaire overall walking ability and 22 complex locomotor skills.

At GIL the data was drawn from all previous clinical patients, while at SLC the data came from a cohort of 350 consecutive individuals identified between February and December of 2019.

### 2.2 GMFCS level and GMFM-66 Score

The GMFCS level for individuals was assessed by an experienced, licensed physical therapist during the visit. For data analysis, individuals were categorized by their initial GMFCS level assignment.

Administering the GMFM-66 is not part of standard practice for routine clinical gait analysis at GIL. However, there were a modest number of individuals in our database for whom GMFM-66 scores were available. These scores had been measured by a licensed physical therapist with training in GMFM testing. The available GMFM-66 scores were included if they were measured within one month of the gait evaluation. At GIL, the average days between collection of FAQ data and GMFM-66 evaluation was 4.7 days. The GMFM-66 is a standard assessment at SLC for children diagnosed with CP. At SLC the GMFM-66 and FAQ were collected on the same day.

### 2.3 Computing the FAQt

All computations were performed in R (9). Linear regression was done using the base R functions. Bayesian modeling was done using Rstan (10)

The FAQt is a weighted sum of overall walking ability and complex locomotor skills that a child can do with relative ease. The FAQt is computed using the following steps:

- **Dichotomize Skills (if necessary)**: At GIL, each of the 22 higher-level skills on the FAQ questionnaire was dichotomized into able or unable. Skills rated as “easy” or “a little hard” were re-coded as able and skills rated as “very hard”, “cannot do”, or “too young” were re-coded as unable. At SLC, the skills were natively reported in dichotomized form.
- **Retrieve existing weights based on existing item difficulties**: Weights for each of the 22 complex locomotor skills and for overall walking ability were derived from the existing item difficulties (6,7). The weights for the FAQt were defined to be the inverse logit of the item difficulties (Table 1).
- **Sum weighted skills to get raw FAQt**: The 23 scores (each of the 22 complex locomotor skills and the overall walking ability) were weighted and summed to form the raw FAQt score:

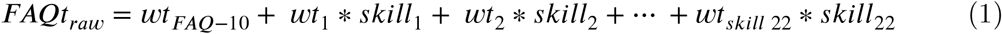 Where ***FAQt***_***raw***_ is the raw (un-scaled) FAQt score, ***wt***_***FAQ***−10_ is the weight for overall walking ability, ***wt***_***k***_ are the weights for the 22 complex locomotor skills, and *skill*_*k*_ takes on the value 1 (able) or 0 (unable).
- **Scale FAQt (0-100 points)**: The maximum total score for an individual with an overall walking ability rated 10, and the ability to perform all skills is 1370. The final (scaled) FAQt score was computed as:

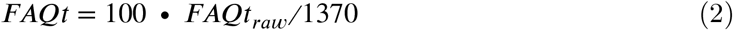

### 2.4 Validation

To test *<u>concurrent validity</u>*, we compared the FAQt to the GMFM-66. We further demonstrated that FAQt evolves with age in a manner like GMFM-66 when stratified by GMFCS level (3). To test *<u>external validity</u>*, we examined whether these relationships differed between the two centers.

**Table 1.** Skill Weights.

| Skill | Weight | Skill | Weight |
| --- | --- | --- | --- |
| Ice/Roller Skate | 94 | Run | 55 |
| Jump Rope | 91 | Kick with Left Foot | 45 |
| Ride 2-Wheel Bike | 88 | Ride 3-Wheel Bike | 45 |
| Hop on Right Foot | 84 | Kick with Right Foot | 44 |
| Hop on Left Foot | 84 | Step over (Lead Left) | 43 |
| Climb Stairs No Rail | 72 | Step over (Lead Right) | 43 |
| Run with Control | 70 | Step Backwards | 40 |
| Ride Escalator Indep. | 67 | Step off of Curb | 40 |
| Jump off Step | 61 | Turn in a Tight Area | 40 |
| Get On/Off Bus | 56 | Walk with Object | 31 |
| Walk with Fragile Obj. | 56 | Climb Stairs Using Rail | 29 |
| **Walking Ability** |  |  | **Weight** |
| Level 10: | Walks, runs, and climbs on level and uneven terrain without difficulty or assistance |  | 94 |
| Level 9: | Walks outside the home for community distances, easily gets around on level ground, curbs, and uneven terrain, but has difficulty or requires minimal assistance with running, climbing and/or stairs |  | 76 |
| Level 8: | Walks outside the home for community distances, is able to perform curbs and uneven terrain in addition to level surfaces, but usually requires minimal assistance or supervision for safety |  | 58 |
| Level 7: | Walks outside the home for community distances, but only on level surfaces (cannot perform curbs, uneven terrain, or stairs without assistance of another person) |  | 41 |
| Level 6: | Walks more than 15-50 feet outside the home, but usually uses a wheelchair or stroller for community distances or in congested areas |  | 27 |
| Level 5: | Walks more than 15-50 feet but only inside at home or school (walks for household distances) |  | 19 |
| Level < 5 |  |  | 0 |

To compare FAQt to GMFM-66 we built a linear model including main effects of GMFM-66 and Center (GIL or SLC), as well as an interaction between GMFM-66 and Center:

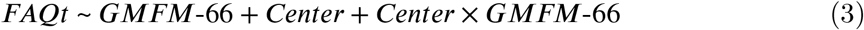

Concurrent validity was measured by the adjusted variance in FAQt accounted for by GMFM-66 (adjusted r^2^). External validity was tested by examining the *Center* and *Center* × *GMFM*-66 terms. Note that if these terms are significant, it implies a lack of external validity, since the center where the data is collected is a meaningful predicter. A final model was defined by testing for significance of terms containing *Center* and retaining only significant terms.

Visual analysis of the FAQt versus age data and the form of the existing GMFM-66 growth curves suggested a generalized growth model (11). Such a model allows for the acquisition of new skills with age, reaching a plateau at locomotor maturity. To examine the evolution of FAQt with age we built a model of the form.

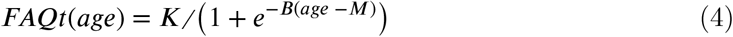

In equation 4, the parameter ***K*** is the FAQt asymptote at maturity, ***B*** is the growth rate, and ***M*** is the age at which the maximum rate of increase in FAQt occurs. We built a full model that allowed for varying effects by GMFCS level, center, and individual. Thus, in the full model, each parameter can be written as (e.g., for K)

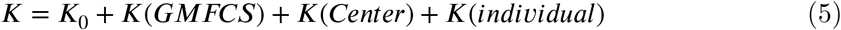

We examined external validity by testing the equality of ***K, B***, and ***M*** across centers. A reduced model was defined retaining significant terms.

## 3 Results

### 3.1 Participant Characteristics

Participants reflected the general referral base of the motion analysis services, with relatively equal numbers of individuals classified as GMFCS I-III (Table 2). The two centers had relatively well-matched percentages of individuals within GMFCS levels, as well as comparable age and sex distributions.

**Table 2.** Participant Characteristics.

|  | GMFCS I |  | GMFCS II |  | GMFCS III |  |
| --- | --- | --- | --- | --- | --- | --- |
|  | GIL | SLC | GIL | SLC | GIL | SLC |
| n (%) | 966 (32) | 126 (43) | 1201 (39) | 122 (42) | 869 (29) | 45 (15) |
| n unique (%) | 577 (34) | 126 (43) | 644 (38) | 121 (42) | 464 (28) | 44 (15) |
| sex = m (%) | 550 (56.9) | 55 (43.7) | 722 (60.1) | 67 (54.9) | 483 (55.6) | 29 (64.4) |
| age (mean (SD)) | 10.3 (3.6) | 9.6 (3.4) | 10.2 (3.8) | 11.4 (3.7) | 9.8 (3.7) | 10.8 (4.1) |
| height (mean (SD)) | 137.1 (20.2) | 134.4 (20.2) | 134.2 (20.8) | 140.1 (18.3) | 128.1 (19.9) | 133.7 (21.2) |
| weight (mean (SD)) | 35.0 (16.0) | 31.9 (15.2) | 34.3 (17.0) | 37.6 (17.4) | 31.1 (14.5) | 35.0 (17.4) |

Note: Age in years, height in cm, weight in kg

### 3.2 FAQt vs. GMFM-66

In the linear model terms that included *Center* were found to be statistically insignificant, with p-values around .80 (Table 3). The reduced model was formed by excluding *Center* terms and re-fitting the model to the data.

**Table 3.** FAQt vs. GMFM-66.

| Term | Estimate | Std. Error | P Value | Model |
| --- | --- | --- | --- | --- |
| **Full Model** |  |  |  |  |
| (Intercept) | -59.59 | 19.84 | 0.00 | Full |
| centerSLC | 4.54 | 20.65 | 0.83 | Full |
| gmfm66 | 1.38 | 0.31 | 0.00 | Full |
| centerSLC:gmfm66 | -0.07 | 0.32 | 0.83 | Full |
| **Reduced Model** |  |  |  |  |
| (Intercept) | -55.41 | 5.15 | 0.00 | Reduced |
| gmfm66 | 1.31 | 0.07 | 0.00 | Reduced |

There was a strong linear relationship between GMFM-66 and FAQt (Figure 1). The GMFM-66 explained 54% of the variance in FAQt (adjusted r^2^), which was slightly higher than the adjusted r^2^ for the full model that included *Center* effects. Remarkably, the x-intercept of 42 is approximately equal to the GMFM-66 score of 46.9 (44.7 – 49) that a person would obtain by scoring maximally on all elements of parts A, B, C (lying, rolling, sitting) of the GMFM-66, and being unable to complete any elements of parts D and E (standing, walking, and running) (12). This supports the premise that the locomotor function embodied in the FAQt is related to domains D and E of the GMFM-66.

**Figure 1.**
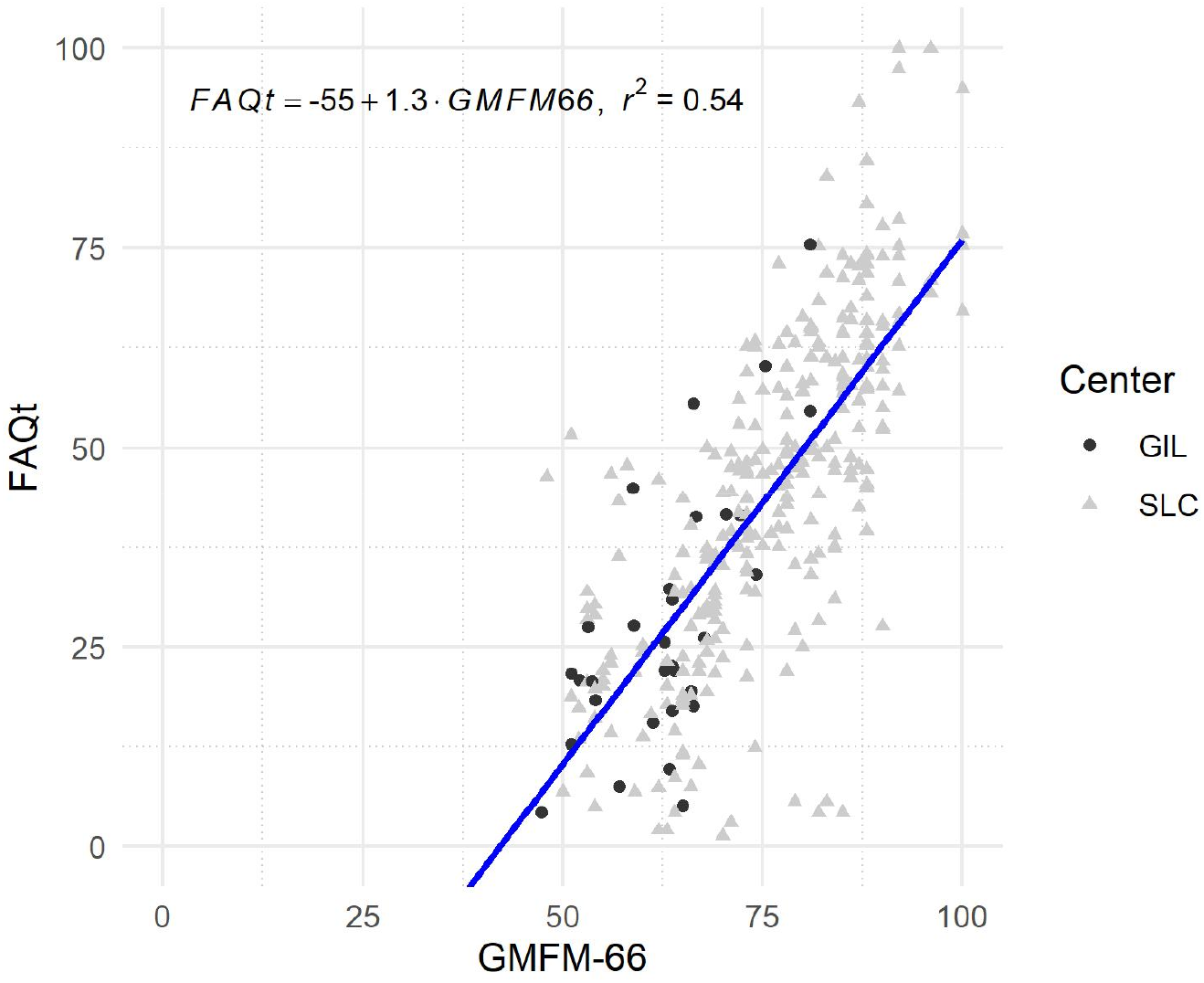
There was a strong linear relationship between GMFM-66 and FAQt. The x-intercept (42) is approximately equal to the GMFM-66 score obtained for completing all tasks on the lying, rolling, and sitting dimensions and none in the standing, walking, and running dimensions (46.9 (44.7 – 49)). The axis limits are the full scale of each measure.

### 3.3 FAQt vs. Age Stratified by GMFCS Level

In the growth model, the effects of center on the parameters ***K, B***, and ***M*** were indistinguishable from zero and from each other. As a result, the center terms were dropped, and a reduced model with only overall and GMFCS level effects was computed. The reduced model of FAQt evolution with age, without center effects, resulted in sensible results (Figure 2). Note that there is longitudinal data from GIL, but lines connecting an individual’s measurements across ages are omitted for clarity.

**Figure 2.**
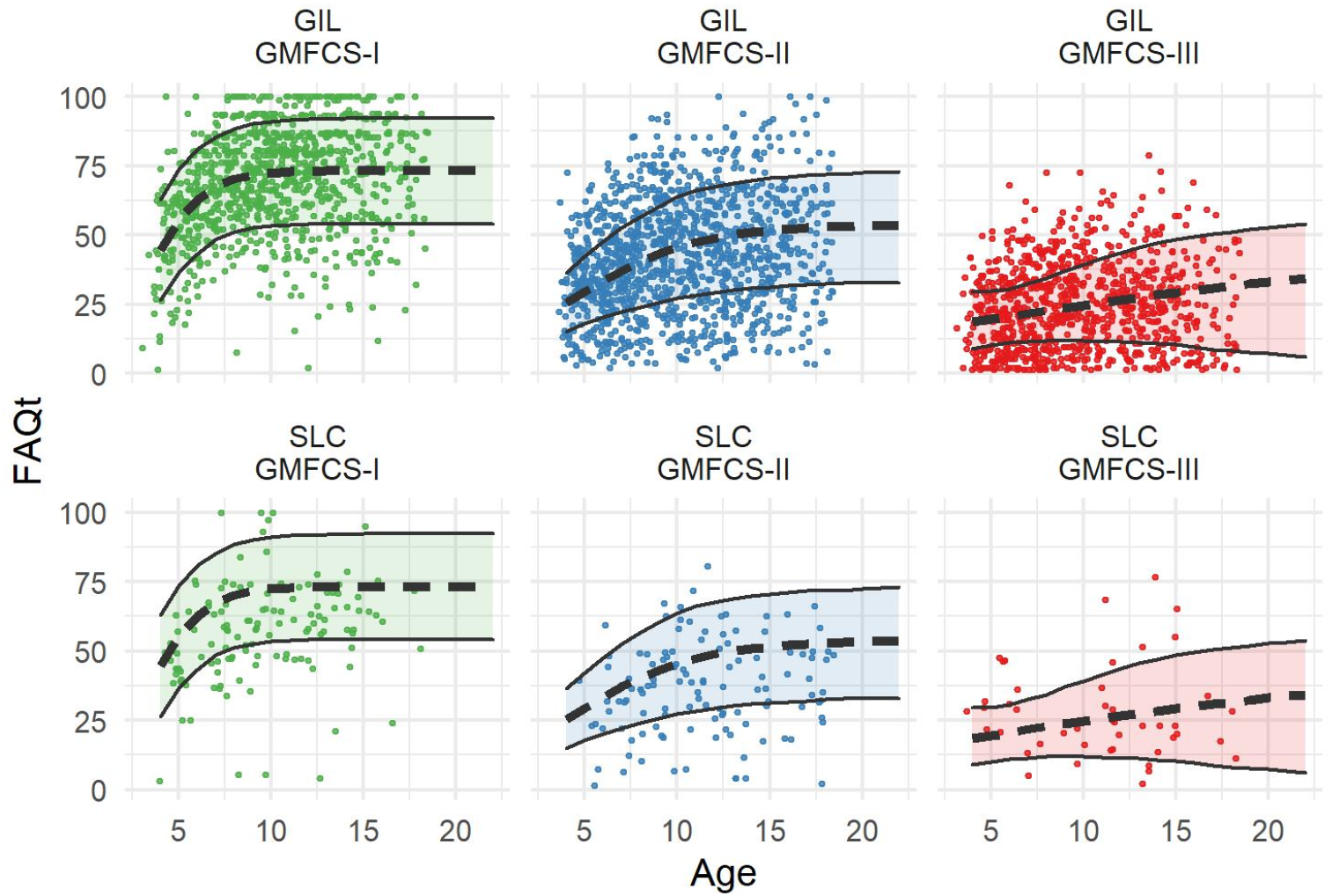
Evolution of FAQt with age, stratified by GMFCS level (columns) and center (rows). Thick dashed lines are the mean response for the given GMFCS level, the shaded area is the 95% credible interval, dots are individual participant trajectories. The FAQt evolves earlier, faster, and to a higher asymptote in higher functioning GMFCS levels. The evolution with age mirrors that of GMFM-66 vs. age stratified by GMFCS level.

The growth in FAQt was faster and reached a higher mature asymptote in higher functioning GMFCS classes (Figure 2). The growth model parameters (asymptote - ***K***, rate of growth - ***B***, and age at maximum growth - ***M***) differed across GMFCS levels (Table 4).

**Table 4.**
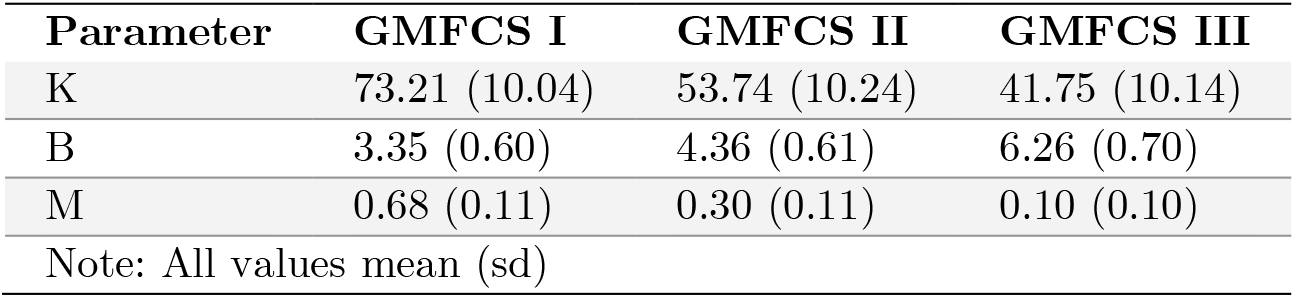
Growth Model Results.

| Parameter | GMFCS I | GMFCS II | GMFCS III |
| --- | --- | --- | --- |
| K | 73.21 (10.04) | 53.74 (10.24) | 41.75 (10.14) |
| B | 3.35 (0.60) | 4.36 (0.61) | 6.26 (0.70) |
| M | 0.68 (0.11) | 0.30 (0.11) | 0.10 (0.10) |
| Note: All values mean (sd) |  |  |  |

### 3.4 Case study

An example of an individual categorized as GMFCS II is provided to clarify the longitudinal nature of the data, and the way individual responses can be fit by the model (Figure 3). The uncertainty bands for future FAQt values of the individual (dotted lines and shaded area) are the 95% credible interval from the posterior distributions. Note that an individual’s expected trajectory and associated credible interval evolve as additional measurements are acquired. In this example, the individual’s predicted FAQt trajectory started out below her GMFCS level peers but ended up almost identical to these peers by age 12.5. Note that each subsequent measurement was near the upper limit of the prediction bands from the previous time point, reflecting an individual who is consistently exceeding model projections.

**Figure 3.**
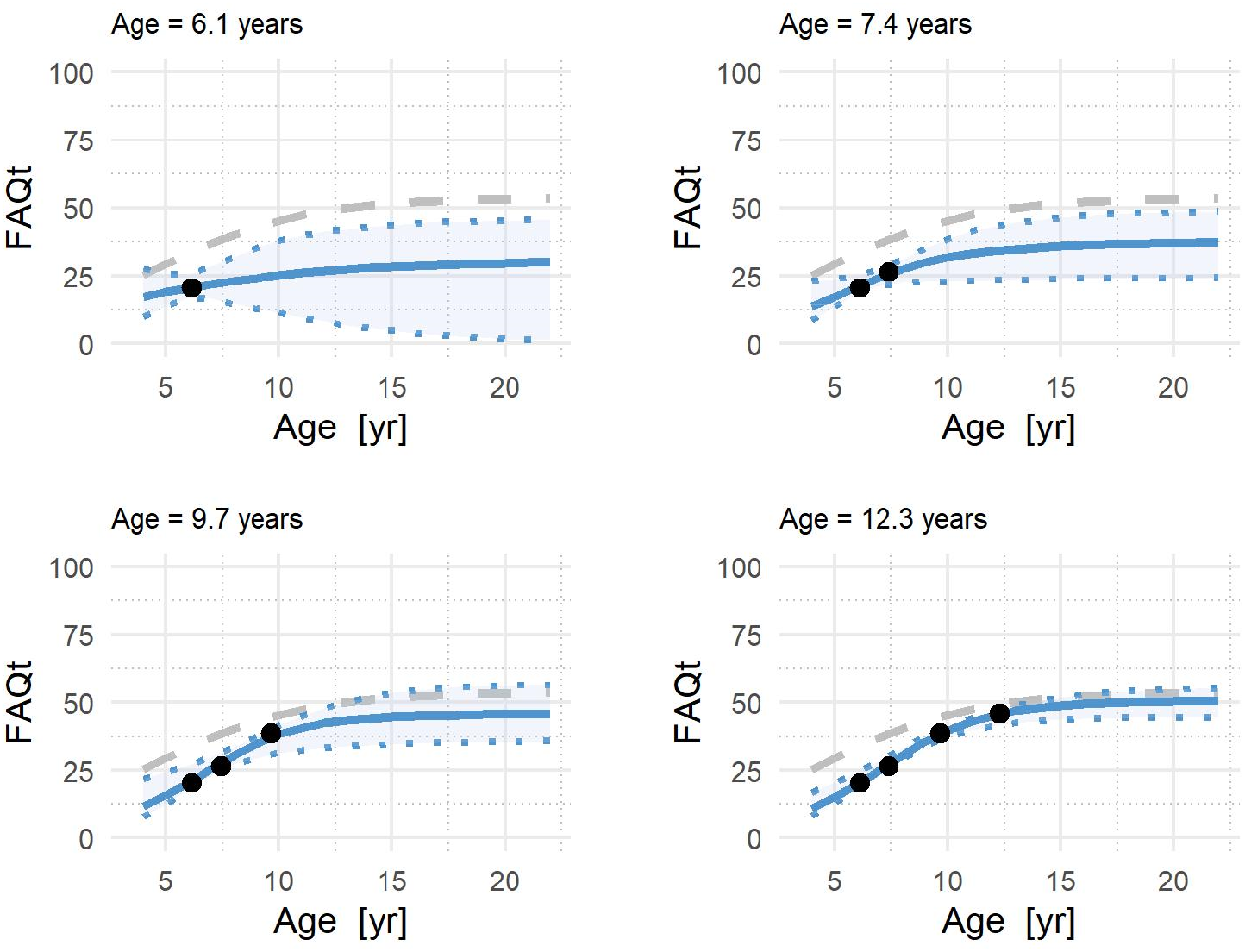
Example of FAQt vs. age trajectories for one individual (GMFCS II) with four responses over time. The dashed line indicates the predicted mean response for GMFCS level II. The solid line is the predicted FAQt vs. age fit for the individual. The dotted lines and shaded area are the 95% credible interval for the individual’s predicted FAQt vs. age trajectory. As more data is acquired, both the predicted future FAQt and its credible interval change.

## 4 Discussion

### 4.1 Validity

The FAQt is proposed as a new measure of locomotor function based on patient reported skills. The FAQt showed strong concurrent validity with respect to the gold-standard GMFM-66 and strong external validity with respect to the two centers in the study. Furthermore, the FAQt is derived from a survey that has previously been shown to have strong content, construct, and concurrent validity, and represent a unidimensional measure of locomotor skill (6).

The FAQt showed a strong linear relationship with GMFM-66. The adjusted r^2^ showed that over half the variance in FAQt was explained by GMFM-66. Furthermore, the model without center effects had a slightly *higher* adjusted r^2^ compared to the model with center effects, supporting the external validity of the FAQt.

The evolution of FAQt with respect to age, stratified by GMFCS level, showed that lower function by GMFCS level led to a lower FAQt at maturity, increased FAQt at a slower rate, and a later age of peak FAQt growth rate. This all matches clinical experience and the widely used motor development curves for CP (13).

The effects of center were found to be insignificant and indistinguishable from zero. This again supports the proposition that the FAQt has external validity, and can be compared across centers (e.g., in multi-center studies).

### 4.2 Limitations

From the standpoint of the measure itself, the primary limitation is the list of activities included in the FAQ questionnaire. These questions span a variety of locomotor skills but are not intended to capture function broadly across activities of daily living. For example, the FAQ does not ask about items related to domains A, B, and C of the GMFM-66 (lying, rolling, and sitting). The result is that, in terms of relation to the GMFM-66, the FAQt largely reflects domains D and E (standing, walking, and running). Furthermore, some of the tasks are either so easy (e.g., climb stairs using a railing, ∼90% can do) or so hard (e.g., ice/roller skating, ∼5% can do) that they are not of much discriminatory value, though they might be useful in a computer-adaptive testing framework.

Another limitation of this study is that we only examined FAQt in children classified as GMFCS I, II, or III. This is due to the ambulatory nature of the referral base at the participating gait laboratories, as well as the skills included in the FAQ questionnaire – almost none of which are doable in children classified as GMFCS IV or V.

The FAQt relies on self-reported abilities. As such they are subjective, and not confirmed by demonstration. People may over- or under-rate their abilities based on complex personal factors. This is a limitation inherent in all patient reported outcomes. In addition, some centers use an inherently dichotomous version of the FAQ questionnaire (SLC) while others use a version that rates skills across four difficulty levels (GIL). In this study, we showed that the two approaches yield statistically and clinically comparable results. While the variability in how the questionnaire is administered is not ideal, it is beyond the scope of this study or these authors to address.

The final important limitation to discuss is the passive nature of the participant enrollment. This study examined retrospective data collected in a clinical laboratory. As such, there are several obvious sources of bias to consider. One is the representativeness of the patient population, which is probably acceptable, since the patients span a wide range of function, as measured by overall walking ability and GMFCS level. More potentially impactful is the role that treatment and follow-up have on our estimates of the evolution of FAQt with age. For one thing, we cannot readily understand how much of the change in function with age is due to maturation and how much is due to treatment. This weakness is also present in the widely used GMFM-66 versus age curves. Additionally, not everyone who gets treated returns for follow-up, and those who do return may result in a biased estimate in the development of function. While this limitation does not affect the relationship between FAQt and GMFM-66, it may affect the evolution of FAQt with age.

### 4.3 Conclusion

We believe the ease of administration, the simple interpretation, and the sensible “growth-curves” included in this study make the FAQt a valuable addition for both clinical assessment and research into locomotor function in children with CP. Much of the content of the FAQt is familiar. Many of the 22 higher level skills are similar (or identical) to skills used in assigning a GMFCS level or computing a GMFM score. Whether due to mimicry, coincidence, or the inherent importance of these skills, the overlap contributes to the high level of concurrence.

The FAQt is not intended to replace the GMFM-66. If time, budget, and logistics permit, the GMFM-66 is an outstanding measure of locomotor function that uses direct patient demonstration of ability. The FAQt is a simple and reasonable alternative in situations where patient report, rather than demonstration, is acceptable, and when the in-person measurement required for the GMFM-66 is not practical. Examples of these situations could include large-scale studies, remote assessment, and longitudinal assessment requiring many measurements on an individual.

## Data Availability

The datasets generated during and/or analysed during the current study are not publicly available due to the fact that they contain protected health information, but may be available from the corresponding author on reasonable request.

## Notes

### Competing Interest Statement

The authors have declared no competing interest.

### Funding Statement

There was no external funding for this study.

